# Real-Time PCR-based diagnosis of human visceral leishmaniasis using urine samples

**DOI:** 10.1101/2022.07.05.22277270

**Authors:** Samiur Rahim, Md. Robed Amin, Mohiuddin Sharif, Mohammad Tariqur Rahman, Muhammad Manjurul Karim

**Affiliations:** Department of Microbiology, University of Dhaka, Dhaka 1000, Bangladesh; Department of Medicine, Dhaka Medical College, Dhaka 1000, Bangladesh; Faculty of Dentistry, University of Malaya, Kuala Lumpur 50603, Malaysia

**Keywords:** Kala-azar, *Leishmania donovani*, Real Time PCR, Urine, Visceral leishmaniasis

## Abstract

**Background:** Diagnosis of visceral leishmaniasis (VL) through the detection of its causative agents namely *Leishmania donovani* and *L. infantum* is traditionally based on Giemsa-stained smears of bone marrow, spleen aspirates, liver or lymph node. Collection of these samples involve invasive procedures and carries the risk of fatal hemorrhage especially during splenic aspiration. Earlier, we reported a Polymerase Chain Reaction (PCR)-based diagnosis of *L. donovani* in peripheral blood using a novel set of PCR primers with absolute specificity (Khatun et al. 2017). Using the same set of primers and PCR conditions, here we describe diagnosis of *L. donovani* from urine, for a non-invasive, rapid and safe diagnosis.

**Methods:** Diagnosis of *L. donovani* was carried out using urine samples collected from clinically diagnosed VL patients (n=23) of Bangladesh in Real Time PCR. Test results were validated by comparing blood samples from the same set of patients. Sensitivity and specificity of this diagnosis was analyzed using retrospective bone marrow samples, collected earlier from confirmed VL patients (n=19) (Khatun *et al*. 2017).

**Results:** The method showed 100% sensitivity in detecting VL in urine and corresponding blood samples and bone marrow samples, as well as 100% specificity in control groups.

**Conclusion:** Urine-based diagnosis could be a patient-friendly, non-invasive approach for VL detection with precision and perfection.

## Introduction

The protozoan parasite *Leishmania donovani* (LD) is the causative agent for visceral leishmaniasis (VL) or kala-azar (KA), which is a vector-borne disease transmitted by the bite of female sand fly. The parasite exists in two stages to complete its life cycle: Amastigote (developed in human) and Promastigote (in sand fly). The amastigote form residing in the cells of mononuclear phagocytic system (MPS), like macrophage, neutrophil and endothelial cells of human, multiplies by binary fission until the MPS cells become enlarged. Infected cells are eventually ruptured and the parasites are liberated in the circulation and invade fresh cells; the cycle is repeated until all cells of MPS are affected. Proliferation of MPS cells leads to massive splenomegaly and hepatomegaly. The bone marrow is also involved, resulting in pancytopenia.

Approximately 90% of the 500,000 estimated annual cases of kala azar occur in rural areas of Bangladesh, India, Nepal, Sudan and Brazil [1] whereas Bangladesh, India and Nepal account for 60% of global cases caused by LD. This disease accounts for 40,000 deaths annually [2]. In Bangladesh, 45 out of 64 districts are endemic for VL, while 20 million people are at risk of developing VL [3].

In recent years, Polymerase Chain Reaction (PCR) as well as Real-Time PCR based diagnostic methods have been evaluated for the diagnosis of VL and shown excellent sensitivities and specificities using spleen aspirate, bone marrow, lymph node and blood [4–7]. These processes, however, are invasive and some of them require skilled health personnel to collect samples. Furthermore, possible risk of internal hemorrhage during collection of splenic aspiration or bone marrow biopsy could be lethal for patients. Urine, on the other hand is a non-invasive specimen, which could be a potential source for detection of VL, for it is a systemic and chronic disease associated with acute kidney injury (AKI).

Multiple cases of proximal tubular injury and glomerular inflammation have been reported to be related with VL associated nephropathy [8–12]. It has been postulated that glomerulonephritis occurs due to deposition of immune-complex in kidney. *Leishmania* bodies in kidney were also detected in light and electron microscopy [13], therefore it is likely to be present in urine.

Given the involvement of VL associated nephropathy, a PCR-based diagnosis of VL caused by *L. infantum* using urine was developed in Brazil [14,15]. However, similar strategy is yet to be adopted in Bangladesh, India and other countries of south Asia and south-east Asia for detection of VL caused by *L. donovani*, a species indigenous in this region [16]. Earlier, this lab developed a blood-based method of diagnosing VL using a novel set of primers, MK1 primer pair specific to *L. donovani* that produced 98% sensitivity and 100% specificity [7]. Here, we report the suitability of the same set of primers [7] using urine as sample from patients with VL in both real time PCR and conventional PCR settings, thereby establishing a patient-friendly, non-invasive method of disease diagnosis.

## Materials and Methods

### Ethical clearance and study group

An Ethical clearance from Dhaka Medical College Hospital (DMCH), Dhaka, Bangladesh (reference: Memo No. ERC-DMC/ECC/2021/409, dated 23 Nov 2021), and Bangladesh Medical Research Council (reference number BMRC/NREC/2013-2016/816, dated 8 May 2014) as well as formal written consents from individuals participated in this study were duly obtained at the onset of the study. For participants under 18 years of age, the formal written consent was obtained from the respective parent/ guardian.

Twenty-three patients with clinically diagnosed VL (CDVL) group were enrolled in this study; twenty-one of them were admitted in the DMCH, while the rest two were in Bangabandhu Sheikh Mujib Medical University (BSMMU). Seventeen of them were adult male, five were adult female and one was a male child. Twenty participants were enrolled as healthy control (HC) group; ten each from VL endemic and non-endemic areas. There were nineteen confirmed VL (CVL) patients, and twenty-five patients formed the disease control (DC) group with symptomatic diseases, i.e., tuberculosis (n=10), malaria (n=3), and dengue (n=12) as reported earlier [7] (first row of Table 2).

### Clinical diagnosis of the patients

Clinical diagnosis of the patients was confirmed by the clinicians at DMCH, and BSMMU, Dhaka, Bangladesh. Patients with clinical signs such as fever, splenomegaly and hepatomegaly symptoms who were admitted in DMCH and BSMMU until March 2022 were confirmed with VL by on-site serology test (rk39-based Immunochromatographic test (ICT)), microscopy of parasitology (observation of amastigote in spleen or bone marrow aspirations), and medical examination by resident clinicians.

### Sample collection

#### Urine

Urine samples were collected from all CDVL (n=23) and HC (n=20) groups. Briefly, 50 mL of urine were collected from each individual in a 100 mL plastic container (Falcon, USA) and were centrifuged at 12,000 *×g* for 5 minutes; then the pellet was suspended in phosphate buffer saline (PBS) for DNA extraction.

#### Blood

2 ml venous blood samples were collected from the same set of CDVL (*n* = 23), HC (n=20), and DC (n = 25) groups. To separate buffy coat from whole blood, blood was centrifuged at 300 rpm for 10 minutes. Supernatant plasma above the buffy coat layer was removed and the layers of buffy coat followed by erythrocytes underneath (to a depth of about 1 mm) were collected using a Pasteur pipette.

#### Urine

Urine samples were collected from all CDVL patients (n=23). Briefly, 50 mL of urine were collected from each individual in a 100 mL plastic container (Falcon, USA) and were centrifuged at 12,000 *×g* for 5 minutes; then the pellet was suspended in phosphate buffer saline (PBS) for DNA extraction. Urine (n=20) was also collected from healthy control (HC) group consisting of healthy individuals of non-endemic (Dhaka, n = 10) and endemic areas (n = 10).

#### Blood

2 ml venous blood samples were collected from the same set of CDVL patients (*n* = 23) from where urine samples were collected. On the other hand, venous blood was also collected from both volunteers of HC group (n=20), and patients with symptomatic diseases similar to VL of disease control group (n = 25; 10, 3 and 12 patients diagnosed for tuberculosis, malaria and dengue respectively). To separate buffy coat from whole blood, blood was centrifuged at 300 rpm for 10 minutes. Supernatant plasma above the buffy coat layer was removed and the layers of buffy coat followed by erythrocytes underneath (to a depth of about 1 mm) were collected using a Pasteur pipette.

#### Bone marrow

Retrospective bone marrow samples collected from the CVL (n=19) patients participated in an earlier study [7] were considered for the current analyses.

#### Positive control

*L. donovani* DNA, obtained from culture positive promastigote was used as the positive control.

### DNA extraction

Total DNA was extracted from urine using QIAamp DNA Mini Kit following manufacturer’s instructions. In order to extract DNA from blood samples, 50μL of blood buffy coat was homogenized with 1 mL of Tween 20, nonidet P-40, NaOH, Tris pH 7.2 (TNNT) buffer and 10μL of a proteinase K solution (containing 320μL of the enzyme/mL) in a 1.5 mL micro-centrifuge tube. Then it was incubated for 3 hours at 56°C followed by an incubation at 72°C for 10 min. The homogenate was then kept at 37°C overnight before 200μL of a phenol–chloroform– isoamyl alcohol mixture (25:24:1 by volume) were added. After vigorous shaking, the mix was centrifuged (10,000 ×g for 10 min), and then the DNA in the aqueous layer was separated, and precipitated with 400μL cold absolute ethanol. The pellet was re-suspended in 50μL double-distilled water before storing at 4°C.

### Real-Time PCR Reaction

Real-time PCR based detection of VL was conducted using the novel MK1F/R primers which were designed to amplify a 102 bp fragment of leishmanial kinetoplast DNA [7] in the ABI PRISM 7000 system (Applied Biosystems). To quantify the parasite load, a standard curve was constructed using DNA extracted from six 10-fold serially diluted *in vitro* cultured promastigote of *L. donovani* corresponding to 10,000 to 0.1 parasite per microliter. For diagnosis, each 20μL reactions included a 5μL sample of DNA, 1μL of 10 μmol/L of each primer MK1F and MK1R[7], 10μL Promega™ GoTaq™ qPCR Master Mix (containing BRYT Green^®^ Dye having spectral properties similar to those of SYBR® Green I), and 3μL of nuclease-free water. The reaction was performed with an initial denaturation step at 95°C for 10 min, followed by 40 cycles of amplification (95°C/30 sec, 60°C/30 sec). Non-template control (NTC) and positive control were included. ABI PRISM software (version 1.1) was used for result analysis. Melt curve pattern for the PCR amplicons was produced using a temperature gradient from 60°C to 95°C. The melt pattern was analyzed for the pick at 82±2°C, the theoretical melting temperature of the 102 bp-long amplicon of MK1 primer pair.

### Sensitivity, Linearity and Reproducibility of Real Time PCR Assay

A known six 10-fold serial dilutions of DNA (1 ng to 10fg/µL) extracted from *in vitro* cultured promastigotes of *L. donovani* corresponding to 10,000 to 0.1 parasites per microliter was used to determine the minimal number of parasites that could be calibrated by the assay. Each reaction contained 1µL of parasite DNA. All dilutions were tested three times. Three replicates of six 10-fold DNA concentrations were tested in a single run for intra-assay validation. For inter assay validation, similar dilutions were made and three independent runs were performed. The coefficient of variation (CV; represented as the ratio of mean to standard deviation, SD) measures the assay’s variability. Melt curve analysis was conducted to evaluate the analytical specificity of real-time PCR products.

### Conventional PCR Reaction

PCR based detection of VL was conducted using the same MK1F/R primers which were designed to amplify a 102 bp fragment of leishmanial kinetoplast DNA [7]. Five microliters of purified DNA extracted in elution buffer (QIAamp DNA Mini Kit) from either blood or urine samples was amplified as described earlier [7]. The PCR products were analyzed by electrophoresis on a 1.2% (w/v) agarose (Thermo Scientific, Mol Bio Grade, USA) gel, containing ethidium bromide (0.5 mg/mL in TBE buffer), 0.04 M Tris-borate and 0.001M EDTA; thereafter photographed under UV illumination using Gel documentation system (Alpha Imager HP System Versatile Gel Imaging, Santa Clara, CA, USA).

### Statistical Analysis

Online tool Vassar Stats was used to calculate the statistical analyses at 95% confidence interval (CI). The sensitivity and the specificity of the primer pairs were calculated according to the following statistical formulas:

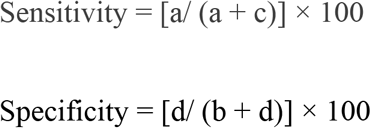

where a, b, c and d represent numbers of true positive, false positive, false negative, and true negative samples respectively.

## Result

### Clinical History of The Patients

The clinically diagnosed VL patients (n= 23) were 16 to 86 years old (median 36 year). They were admitted with a history of 1 to 2.5 months (median 1.6 month) of fever before hospitalization. 72% of the patients had splenomegaly and 63% of them had hepatomegaly while all of them were reported to have anemia as well as weight loss. There was no post kala azar dermal leishmaniasis (PKDL) case in the study. Again, no patients took anti-leishmanial drug during the episode of VL. Samples were collected from CDVL patients admitted either at DMCH or BSMMU. The respective institutions used their standard protocol for the diagnosis of VL in CDVL patients depending on the condition of the patients, for example, microscopic examination of bone marrow (n=1) and splenic aspirate (n=3), and rk39 ICT test (n=19) in addition to the on-site medical examination by the physicians. The subsequent Real Time PCR-based molecular detection was thereafter conducted using urine and blood of the clinically diagnosed VL patients.

On the other hand, DNA extracted from bone marrow samples (n = 19), collected from CVL patients participated in a preceding study [7] preserved in DNA bank of Fermentation and Enzyme Biotechnology Laboratory (FEBL), Department of Microbiology, University of Dhaka were taken into consideration for diagnosis by Real Time PCR.

### Real-Time PCR

#### Analytical sensitivity, linearity, reproducibility of Real time PCR assay

With discernible amplification and melt curve around 81°C, the real-time PCR assay detected as little as 10 fg of LD genomic DNA per reaction, which corresponds to 0.1 parasite (Fig 1). The standard curve produced by gradual dilution of parasite DNA over a 6-log range and cyclic threshold (Ct) values formed a linear relation, with an R^2^ of 0.994 and PCR efficiency of 94% (Fig 1B). The absence of Ct values in the non-template control validated the specific response of the reaction mix. For six distinct concentrations, the intra assay coefficient of variation (CV) of Ct values were, 0.03%, 0.0575%, 0.1222%, 0.1667%, 0.1228% and 1.14% respectively (Table 1). More variability was observed with smaller amount of parasite DNA. Inter assay variation of Ct values for the same dilution series in two independent runs was used to test the assay’s reproducibility. The coefficients of variation between assays were determined to be 1.672%, 1.645%, 2.371%, 2.576%, 1.735% and 3.4% percent, showing strong reproducibility (Table 1).

**Figure 1:**
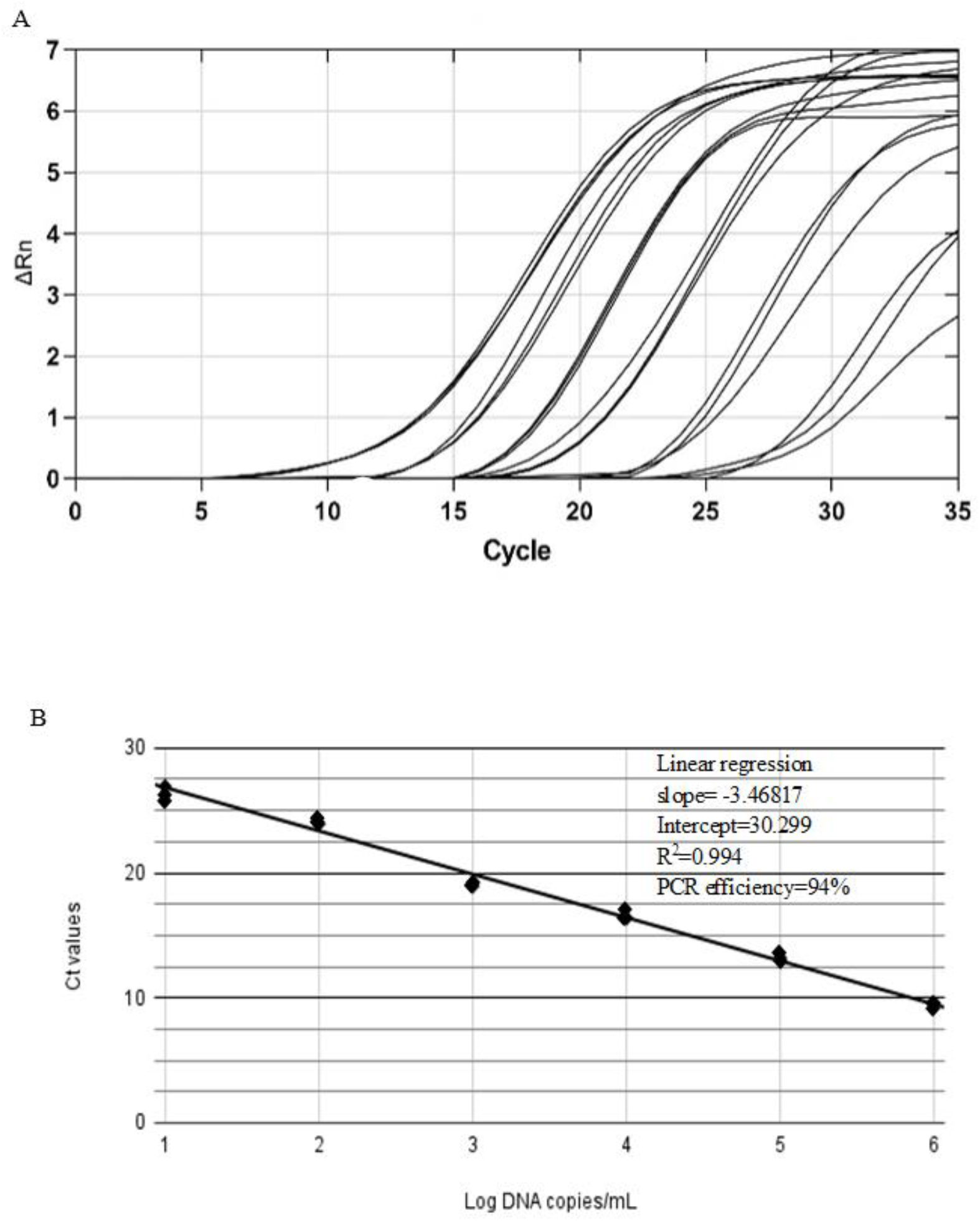
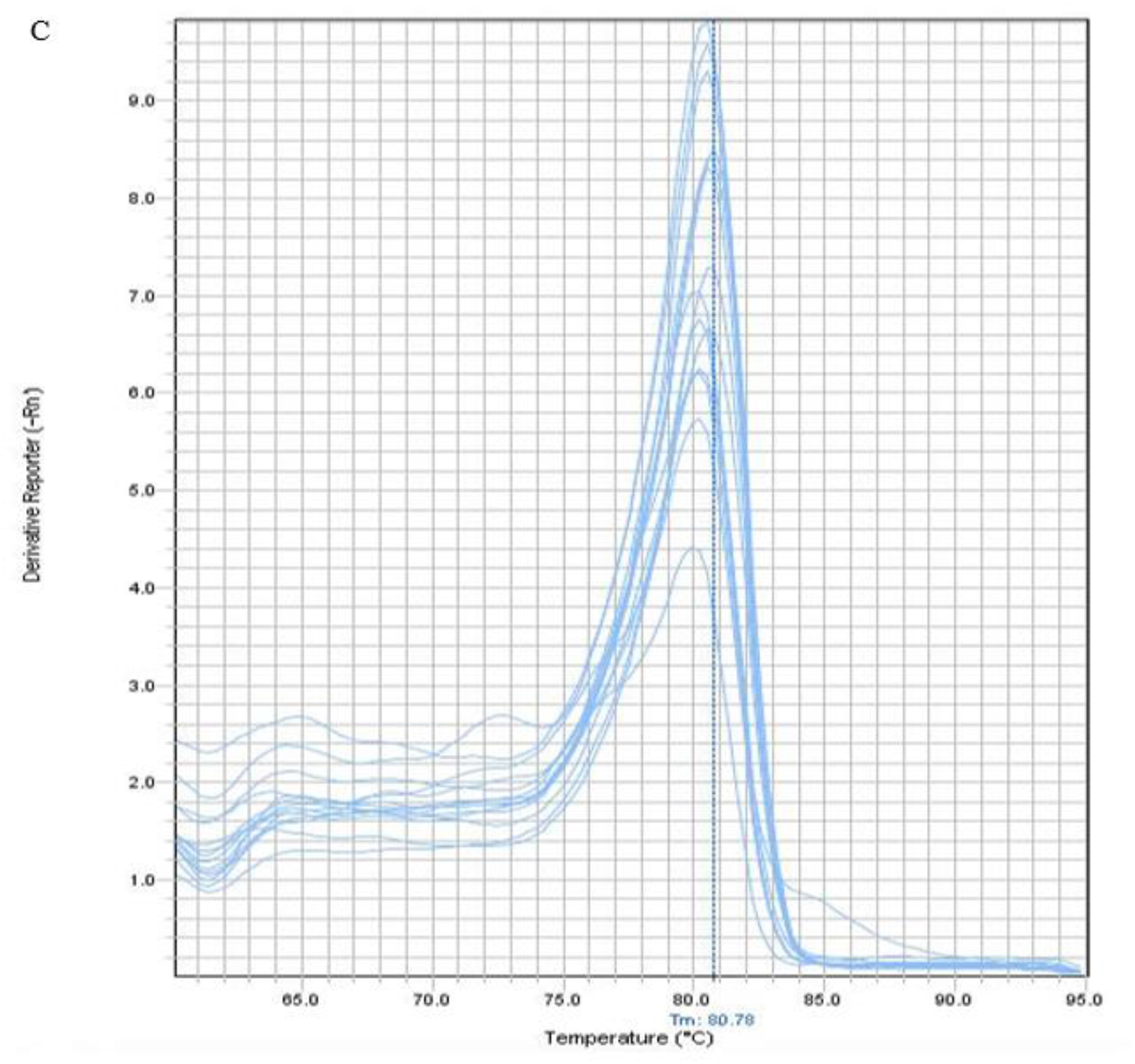
Analytical sensitivity and detection limit of VL diagnosis using Real Time PCR. Amplification curves are shown for DNA extracted from 6 log serially diluted *L. donovani* culture, ranging from 1×10^4^ to 1×10^−1^ parasite per microliter (A). A standard curve is constructed based on the mean of Ct values plotted against serial dilutions containing corresponding *L. donovani* DNA (B). Each point represents the Ct value of an individual sample and the plot represents a linear function (R^2^=0.994). Melt curve of amplification product showing peak near 80.78°C (C).

**Table 1:** Reproducibility of Real time PCR based diagnosis of VL

| Parasite Load<br>(parasite/μL) | Intra assay Variation of Ct value | | | | | Inter assay variation of Ct value | | | |
| --- | --- | --- | --- | --- | --- | --- | --- | --- | --- |
| | Replicate 1 | Replicate 2 | Replicate<br>3 | Mean±SD | CV% | Assay 1 | Assay 2 | Mean±SD | CV% |
| $1 \times 10^4$ | 9.58 | 9.52 | 9.52 | 9.54±0.03 | 0.03% | 9.54 | 9.23 | 9.38±0.157 | 1.67% |
| $1 \times 10^3$ | 13.44 | 13.43 | 13.42 | 13.43±0.008 | 0.058% | 13.43 | 12.99 | 13.21±0.217 | 1.65% |
| $1 \times 10^2$ | 16.39 | 16.35 | 16.35 | 16.36±0.02 | 0.12% | 16.36 | 17.16 | 16.76±0.3975 | 2.37% |
| $1 \times 10^1$ | 18.99 | 18.94 | 18.92 | 18.95±0.031 | 0.17% | 18.95 | 19.95 | 19.45±0.501 | 2.57% |
| $1 \times 10^0$ | 23.9 | 23.98 | 23.91 | 23.93±0.029 | 0.1% | 23.93 | 23.12 | 23.53±0.4083 | 1.73% |
| $1 \times 10^{-1}$ | 26.91 | 27.52 | 26.82 | 27.08±0.31 | 1.14% | 27.08 | 27.99 | 28.03±0.953 | 3.4% |

### Clinical Sensitivity and specificity of Real time PCR based diagnosis of VL

Presence of *L. donovani* was evaluated by determination of the Ct and estimation of melting temperature (Tm) using Real-time PCR method. Here, DNAs collected both from the urine of all of the 23 clinically-confirmed patients produced amplification with MK1F/R primers rendering sensitivity 100% (95% Cl, 85.19-100%) (Table 2). Detectable amplifications were found with Ct values between 15 to 28 (Fig. 2A). This finding was no different when DNAs, collected from buffy coat preparations of blood specimens from corresponding patients were utilized. Retrospective DNAs extracted from bone marrow of 19 VL-confirmed patients also produced amplification with Ct values between 9 to 22, producing an absolute sensitivity, 100% (95% Cl, 82.35-100%) (Fig. 2A). The detected Tm values of all types of samples was near 81°C (Fig. 1C) which is close to the theoretical melting temperature of the 102 bp amplicon, 82°C, therefore validating the specificity.

**Table 2:** Comparative efficiency of Real Time PCR and conventional PCR based assays to diagnose VL using urine, blood buffy coat and bone marrow of CDVL and CVL patients

| Assay | Sensitivity (%), 95% CI |  |  | Specificity (%), 95% CI |  |  |
| --- | --- | --- | --- | --- | --- | --- |
|  | CDVL<br>(n=23) |  | CVL (n=19) | VL negative control participants (n=45) |  |  |
|  | Urine<br>(n=23) | Blood buffy coat<br>(n=23) | Bone marrow<br>(n=19) | Urine | Blood buffy coat |  |
|  |  |  |  | Healthy control (n=20;<br>endemic control n=10; non<br>endemic control n=10) | Healthy control (n=20;<br>endemic control n=10; non<br>endemic control n=10) | Disease<br>control<br>(n=25) |
| Real-Time<br>PCR | 23/23(100%),<br>(85.19-100%) | 23/23(100%),<br>(85.19-100%) | 19/19(100%),<br>(82.35-100%) | 0/20 (100%), (83.16-100%) | 0/20 (100%), (83.16-100%) | 0/25 (100%),<br>(86.28-100%) |
| Conventional<br>PCR | 23/23(100%),<br>(85.19-100%) | 23/23(100%),<br>(85.19-100%) | 19/19(100%),<br>(82.35-100%) | 0/20 (100%), (83.16-100%) | 0/20 (100%), (83.16-100%) | 0/25 (100%),<br>(86.28-100%) |

**Figure 2:**
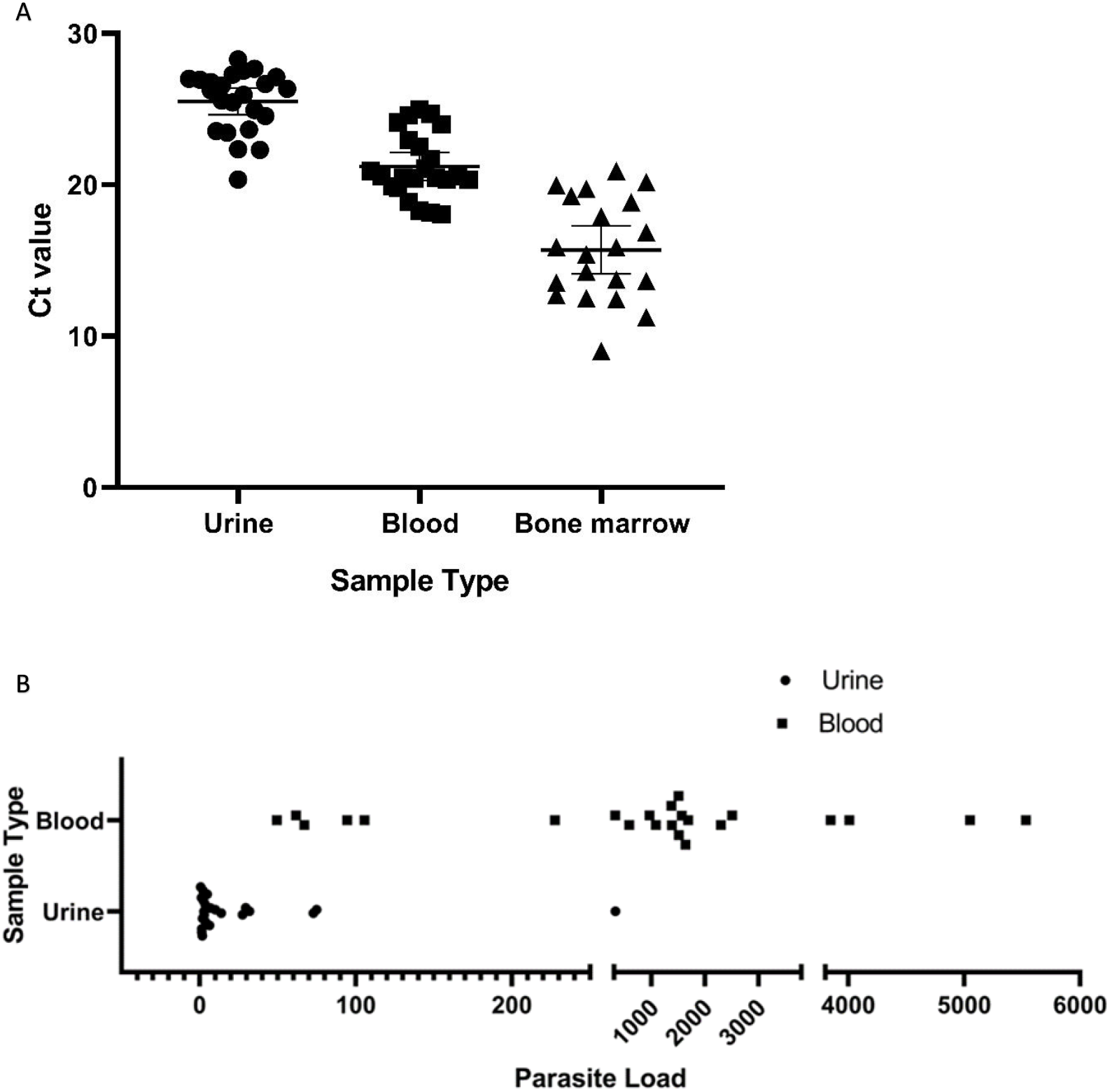
Ct value in CDVL and CVL patients and parasite load in CDVL patients. Ct value in urine from CDVL (n=23), in blood from respective CDVL (n=23) and in bone marrow from CVL (n=19) patients, each point indicates data collected from individual patient (A). Parasite burden in urine and blood samples of CDVL (n=23) patients are illustrated (B).

Parasite load in two different biological samples of CDVL patients was compared and it was found that parasite load in urine was comparatively lower than blood ranging from 1 promastigote/mL of urine in patient-6 to 329 promastigote/mL of urine in patient-11, while blood had a higher load starting from 49 promastigote/mL of blood in patient-8 to as high as 5,532 promastigote/mL of blood in patient-2 (Fig. 2B).

No amplification was observed in both blood and urine samples of individuals, collected from all endemic and non-endemic healthy control with undetected Ct value in real time PCR. Again, no cross reaction was observed with DNA extracted from blood of malaria, tuberculosis and dengue patients indicating 100% (95% Cl, 82.35-100%) specificity of this diagnosis (Table 2).

### Conventional PCR

DNAs collected from the urine specimens of the 23 clinically diagnosed patients, produced the amplicon of 102 bp in polymerase chain reactions when probed with MK1F/R primers (Fig. 3).

**Figure 3:**
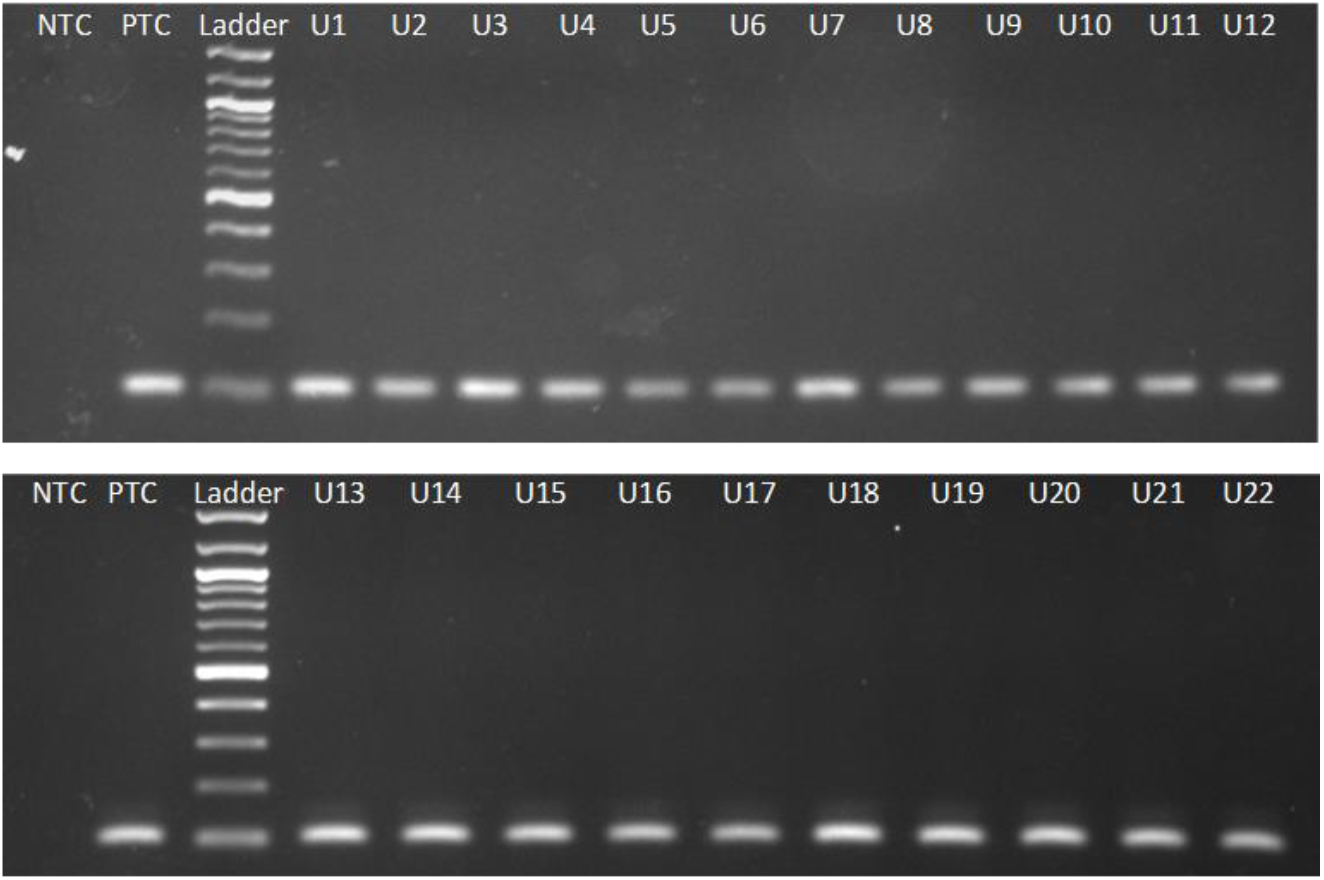
The PCR-amplified bands using novel MK1F/R primers of 102 bp fragment of leishmanial kinetoplast DNA of CDVL patients. Amplicons were separated on a 2% agarose gel generated from 22 urine samples of CDVL patients labeled U1 through U22; PTC was positive template control containing DNA of cultured LD; NTC was negative template control.

The finding was no different when DNAs collected from the buffy coat preparations of the corresponding patients were used as template in a similar set of PCR reaction. The diagnostic sensitivity of VL using urine as specimens was measured 100% (95% Cl, 85.19-100%) when compared to conventionally used specimen and the peripheral blood samples (Table 2). Importantly, none of the 20 control urine samples collected from healthy individuals produced the amplicon, indicating 100% specificity of the analyses (Table 2).

## Discussion

Visceral leishmaniasis is one of the major health issues of tropical and sub-tropical countries like Bangladesh, India, Nepal, Brazil and Iran. However, rapid and accurate diagnosis of the agent is a major concern in controlling this disease. The microscopic detection of amastigote of *Leishmania donovani in* aspirates from lymph node, bone marrow and spleen are the gold standard for the diagnosis, and hold a sensitivity of 53%–65%, 53%–86% and 93%–99% respectively [17]. Although the sensitivity of the bone marrow/ splenic smear is high, collection of these specimens is not only painful but also poses a threat of fatal hemorrhage for the patients.

Alternative serodiagnosis of leishmaniasis (based on antibody detection) including ELISA showed different sensitivity and specificity depending on the type, source, and purity of the antigens. Some *Leishmania* antigens have shown cross reactivity with antigens of other microorganisms [17–19]. However, detection of antibody (IgG) in urine of patients has been used for the VL diagnosis with a sensitivity and specificity up to 98% and 95% respectively, with notable limitations: such as, positive reactions in asymptomatic patients, and inability to distinguish between current, subclinical, or past infections [20, 21].

Polymerase chain reaction (PCR)-based diagnostic methods for the detection of VL have shown excellent sensitivities and specificities using various biological samples: spleen aspirate, blood, lymph node and bone marrow [22–25]. A blood-based method of diagnosis of VL using novel primer MK1F/R specific for *L. donovani* showed 98% sensitivity and 100% specificity [7]. Different studies showed that SYBR green Real time PCR-based diagnosis of VL targeting its minicircle DNA demonstrated sensitivity of 79% to 95.7% in blood samples [26,27]. On the other hand, TaqMan probe based real time PCR has showed up to 100% sensitivity in assays targeting genomic DNA [28].

The use of urine as specimens has gained usefulness because of its non-invasive nature, minimal risk of blood-borne infections, and easy collection method, especially when infants are concerned. Urine-based direct anti-globulin test with the antibody (sensitivity 90.7%), and urine ELISA (sensitivity 93.3%) were reported as useful alternatives [29,30]. Here, we compared a real time PCR-based diagnosis of VL caused by *L. donovani* using urine with that of blood and bone marrow. Primarily, the diagnosis of VL was supported by (1) the observation of amastigote in bone marrow aspirations, (2) response of rk39-ICT in serology, and (3) clinical examination by resident physicians, thereby confirming the diagnosis. In order to develop a PCR-based molecular detection method for VL that would ensure rapidity and precision of the diagnosis, urine samples of the participating patients were used as specimen; there they produced (a) the amplification with Ct values ranging 18 to 28 in a RT-PCR (Fig 2A) in the RT-PCR, and (b) bands of interest with a size of 102 bp in the electrophoretogram in conventional PCR (Fig. 3) after necessary extraction of DNAs. Detectable amplification was also present when analyzing the blood samples of corresponding patients, and in the retrospective 19 bone marrow samples of confirmed VL patients in the Real-Time as well as conventional PCR settings (Table 2)

Regardless of variability in samples, Real Time PCR-based assay is more sensitive than other molecular diagnostics such as conventional PCR or nested PCR. In this study, Real Time PCR assay was able to detect 0.1 parasite per reaction with a detectable melt curve. The limit of detection of this analysis is comparable and, in some cases, better than other currently available dye and probe-based Real Time PCR, while most of them were studied for VL diagnosis from blood or other more invasive biological samples [27,28,31,32]. It is noticeable that the diagnosis using urine showed 100% sensitivity which is same as it showed using blood and bone marrow specimens.

The road map of World Health Organization (WHO) for neglected tropical diseases (NTD) aims to eradicate VL including PKDL from 75 endemic countries by 2030 by emphasizing (a) research on the relationship among the human, vector, parasite and reservoir, (b) development of more specific and user-friendly diagnostic test, and (c) introduction of faster, more effective, safer, cost effective treatment for the VL patients all over the world [33]. Due to its super-sensitive detection limit as little as 10 fg DNA, and absolute level of sensitivity and specificity for urine samples, Real-Time PCR-based diagnosis of VL using MK1 primer can be a suitable detection method to replace blood or more invasive specimens such as bone marrow or spleen. Furthermore, when compared to conventional and nested PCR-based techniques, the ease of one-step molecular detection of RT-PCR reduces work load for the pathologists, thereby fulfilling the patient-friendly diagnostic requirement to achieve the WHO objective. This method can also be used in assessing the response to therapy, monitoring parasite kinetics, infection dynamics and epidemiological survey.

The cohort of this study for diagnosis of VL from urine samples was small (n=23), therefore further study is necessary to ascertain the degree of sensitivity and specificity of the diagnostic procedure. Moreover, the average admittance of the studied patients in the hospital was 1.7 months after the onset of illness (range 1 to 2.5 months). Therefore, the length of this diagnostic strategy beyond 2.5 months’ post-infection needs to be assessed. Nevertheless, this is the first report of its kind in Indian sub-continent to use urine as a clinical specimen for detection of *L. donovani* and its impact in global public health in eradicating this NTD is significant.

## Conclusion

Invasive specimens like bone marrow, splenic aspiration or blood could be substituted by non-invasive specimen, urine to score the infection with *L. donovani* in patients by Real Time PCR without compromising the diagnostic efficiency of VL in Bangladesh, and this success is worth investigating in other VL endemic areas of the world.

## Data Availability

All data and related metadata underlying the findings reported in the submitted manuscript are already provided to be deposited in a public repository, 'medRxiv' as part of the submitted article.

## Acknowledgements

We are thankful to Professor Dr Anowara Begum and Dr Zenat Zebin Hossain, Department of Microbiology, University of Dhaka for providing technical support to conduct Real-Time PCR analysis. We thankfully acknowledge the financial support provided by the University Grants Commission Bangladesh through the University of Dhaka [Regi: Admin-3/12247, dated 16 Sep 2021] in conducting the research.

## Notes

### Competing Interest Statement

The authors have declared no competing interest.

### Funding Statement

This study was supported financially by the University Grants Commission Bangladesh through the University of Dhaka [Regi: Admin- 3/12247, dated 16 Sep 2021]

### Author Declarations

An Ethical clearance from Dhaka Medical College Hospital (DMCH), Dhaka, Bangladesh (reference: Memo No. ERC-DMC/ECC/2021/409, dated 23 Nov 2021), and Bangladesh Medical Research Council (reference number BMRC/NREC/2013-2016/816, dated 8 May 2014) were obtained. In addition formal written consents from individuals participated in this study were duly obtained at the onset of the study. For participants under 18 years of age, the formal written consent was obtained from the respective parent/ guardian.

